# Assessing Type 2 Diabetes and GLP-1 agonist response trajectories with a proteomic atlas of glycemic stages

**DOI:** 10.1101/2025.09.26.25336579

**Authors:** Sivateja Tangirala, Shakson Isaac, Youssef Gehad, Filomene Roquefort, Pauline Gabrieli, Aaron Leong, Josep Mercader, Magda Sevilla-Gonzalez, Federico Baldini, Gary W Miller, Vidhu Thaker, Braden T Tierney, Chirag J Patel

## Abstract

Type 2 diabetes (T2D) progresses through heterogeneous pathways that glycemic staging alone does not resolve. We constructed the Metabolic Atlas of the Proteome in Diabetes (MAP-D), leveraging Olink measurements of 2,923 circulating proteins in ∼42,000 UK Biobank participants to map associations with three cardiometabolic hallmarks - adiposity (BMI), a proxy for insulin resistance (triglyceride-to-HDL cholesterol ratio), and glycemia (HbA1c) - across normoglycemia, prediabetes, and incident T2D. We triangulated cross-sectional associations with bidirectional Mendelian randomization and semaglutide trial proteomics to infer causal directionality. This revealed three distinct causal architectures: adiposity predominantly reshapes the proteome, glycemia is driven by upstream proteins, and insulin resistance shows bidirectional feedback. Integration with trial data identified proteins reversed by therapy and a subset of persistent proteins that remain dysregulated despite GLP-1RA treatment. These persistent proteins are associated with incident coronary artery disease and overlap with targets of approved therapies, nominating candidates for combination strategies beyond GLP-1RA monotherapy.

**Highlights:**

- MAP-D maps 2,923 proteins across three hallmarks and glycemic stages
- Adiposity reshapes the proteome; glycemia is driven by upstream proteins
- Persistent proteins remain dysregulated despite GLP-1RA treatment
- Persistent proteins predict incident CAD and are druggable targets

## Introduction

The stages that are defined by normoglycemia, prediabetes, and type 2 diabetes (T2D) are biologically heterogeneous. Its core clinical hallmarks, including adiposity, insulin resistance, hyperglycemia, and beta-cell dysfunction, arise and contribute unequally to disease onset and shaping variable treatment response ^1–3^. This heterogeneity is only partially captured by current clinical thresholds used to stage disease, motivating molecular resolution of pathways that distinguish normoglycemia, prediabetes, and type 2 diabetes across cardiometabolic hallmarks that are central to risk of complications^4^.

Large-scale GWAS of T2D have highlighted heterogeneous genetic architecture across glycemia, obesity, and insulin resistance^2,5–7,8,9^, with genetic drivers of insulin production (beta-cell function) showing stronger disease associations than insulin sensitivity counterparts, suggesting multiple overlapping biological pathways underlie each hallmark. In contrast, multi-omic approaches (e.g., metabolomics^10^) have mapped pathways and detect perturbations long before diabetes onset^11^. However, existing proteomic studies have not achieved comparable resolution: most pool normoglycemia, prediabetes, and T2D under a single disease construct rather than systematically examining associations with adiposity, insulin resistance, and glycemia across glycemic stages^12–14^.

A glycemic stage-resolved proteomic map is now feasible, given the convergence of biobank-scale proteomic data and new interventional proteomic datasets from GLP-1 receptor agonist (GLP-1RA) trials. Yet it remains unclear how proteins correlated with disease relate to those that change under therapy; clarity in this regard could illuminate mechanisms of risk reduction and anticipate off-target benefits or adverse events.

A central challenge is that much of biobank proteomic data are cross-sectional, precluding direct observation of within-individual disease trajectories. Here, we integrate cross-sectional proteomic associations with bidirectional Mendelian randomization (MR) to establish causal directionality, and compare these relationships against longitudinal proteomic changes from randomized controlled trials (STEP 1 and STEP 2) of semaglutide ^15,16^. Together, these independent lines of evidence allow us to “triangulate” ^17^ proteomic signals, identifying proteins whose hallmark associations, genetic causal estimates, and treatment responses converge to support a role in disease stages.

Here, we pursued three aims. First, we characterized stage-specific proteomic signatures associated with hallmarks of diabetes, including adiposity (BMI), a proxy for insulin resistance (triglyceride/HDL cholesterol (TG/HDL) ratio), and glycemia (HbA1c) across normoglycemia, prediabetes, and incident T2D. Second, we tested whether these hallmarks differ in their causal relationship with the proteome, distinguishing hallmarks that predominantly reshape protein levels from those driven by upstream proteomic pathways. Third, we identified proteins whose hallmark associations are reversed by semaglutide and, conversely, therapeutically persistent proteins that remain dysregulated despite treatment and may signal residual cardiometabolic risk.

## Results

We created an atlas of hallmark-proteomic relationships within three cross-sectional strata defined by glycemia, including normoglycemia, prediabetes, and incident type 2 diabetes (Figure 1A); triangulated these associations against proteomic changes induced by the GLP1-receptor agonist (GLP-1RA) semaglutide in the STEP1/2 trials ^15^ (Figure 1B); used a split-sample bidirectional Mendelian Randomization (MR) to resolve causal direction (Figure 1C); and tested whether persistent proteins (e.g., those whose hallmark association in UKB is not reversed by semaglutide) predicted incident cardiovascular disease in the UK Biobank Olink proteomics subsample (Figure 1D). A summary of the modeling approaches, data, and inclusion criteria used are in Data S1.

**Figure 1.**
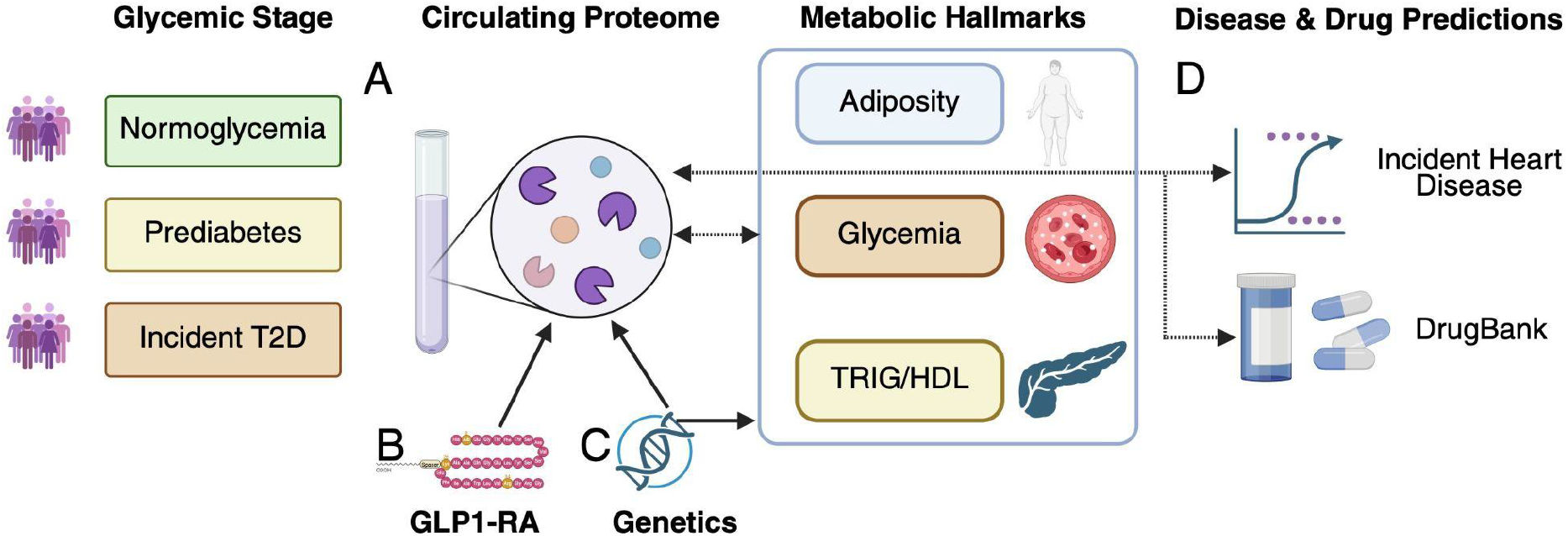
Derivation of the MAP-D Atlas directionality of relationships. A.) within each cross-sectional stage of glycemia (inclusion criteria), we associated the circulating proteome with 3 metabolic hallmarks of diabetes (e.g., BMI, HbA1c, and TG/HDL) in both directions (dashed line). B.) STEP1/2 trials evaluated protein signatures associated with GLP-1RA, and we mapped proteins identified in each glycemic stage (A) onto proteins identified in the STEP RCTs. C.) We used Mendelian randomization to test the causative effects of genetically predicted levels of proteins in B.) and C.) on the hallmarks and of genetically predicted hallmarks on protein levels. D.) We connected proteins with incident heart disease in UK Biobank and found drug targets for those proteins.

### An atlas of protein pathways associated with the trajectory from normoglycemia to type 2 diabetes

We constructed the Metabolic Atlas of the Proteome in Diabetes (MAP-D) (Figure 1), computing cross-sectional associations between 2,923 circulating proteins and hallmarks of T2D, including adiposity (BMI), insulin resistance (TG/HDL ratio), glycemic control (HbA1c) in populations stratified by glycemic stage: normoglycemia (HbA1c <5.7%), prediabetes (HbA1c 5.7-6.4%), and incident T2D (new diagnosis after baseline; see Methods) (Figures 1A, 2-3).

Since proteins and hallmarks were measured concurrently (cross-sectional), regression alone cannot distinguish whether a protein drives a hallmark or responds to it. We therefore performed bidirectional protein-wide association analyses. The primary direction modeled each protein as the outcome and each hallmark as the exposure (Protein ∼ Hallmark; "hallmark-first"), yielding betas that represent the difference in protein level per standard deviation of the hallmark - similar in scale as semaglutide trial effects, enhancing comparability with STEP 1/2 data downstream. We also computed the complementary protein-first associations (Hallmark ∼ Protein), which identify proteins whose levels may causatively influence each hallmark (Figure 1A). All regressions relating hallmarks to proteins were performed in each glycemic group separately (and adjusted for age, sex, assessment center, household income, fasting time, and 40 genetic principal components), resulting in 26,307 total tests in the primary (hallmark-first) direction (2,923 proteins x 3 hallmarks x 3 strata; see Methods).

Among 2,923 proteins measured in the 3 glycemic staged populations, we identified 10,582 Bonferroni-significant (corresponding to a p-value threshold of 1.9×10^-6^ (0.05 / 26,307) “hallmark-first” (modeling protein as the outcome and hallmark as the exposure; hallmark → protein) associations across hallmarks (BMI, HbA1c, and TG/HDL ratio) (5,769 in normoglycemia (n = 39,647 participants), 3,385 in prediabetes (n=5,784), 1,428 in T2D (n=1,369) - a pattern consistent with the declining sample sizes at higher glycemic stages)). In normoglycemia, the stratum with the most Bonferroni-significant hallmark-first associations was TG/HDL (2,163 proteins), followed by BMI (2,042), and HbA1c (1,564) (Table 2). The full Metabolic Atlas of the Proteome in Diabetes (MAP-D) is available at http://mapd.bio and mirrored at https://chiragjp-mapd.share.connect.posit.cloud.

**Table 1.** Cohort description. Demographic and clinical characteristics of UK Biobank participants stratified by glycemic status. Values are mean (SD) for continuous variables and n (%) for categorical variables.

|  |  | **Normoglycemic** | **Prediabetes** | **Diabetes** |
| --- | --- | --- | --- | --- |
| **Total N** |  | 361200 | 51286 | 12068 |
| **Age (years) [mean (SD)]** |  | 55.97 (8.11) | 60.09 (6.69) | 59.84 (7.04) |
| **Sex (%)** | **Female (%)** | 200250 (55.4) | 28440 (55.5) | 4842 (40.1) |
|  | **Male (%)** | 160950 (44.6) | 22846 (44.5) | 7226 (59.9) |
| **Family income (Euros)** | **Less than 18,000 (%)** | 62352 (19.9) | 12941 (30.3) | 3608 (36.8) |
|  | **18,000 to 30,999 (%)** | 77401 (24.7) | 12400 (29.0) | 2746 (28.0) |
|  | **31,000 to 51,999 (%)** | 85260 (27.2) | 9997 (23.4) | 2060 (21.0) |
|  | **52,000 to 100,000 (%)** | 69691 (22.2) | 5969 (14.0) | 1174 (12.0) |
|  | **Greater than 100,000 (%)** | 18906 (6.0) | 1456 (3.4) | 215 (2.2) |
| **BMI (kg/m2) [mean (SD)]** |  | 26.84 (4.37) | 28.75 (5.17) | 31.75 (5.71) |
| **HbA1c (mmol/mol) [mean (SD)]** |  | 34.01 (2.97) | 40.93 (1.83) | 45.80 (12.32) |
| **HDL (mmol/L) [mean (SD)]** |  | 1.48 (0.38) | 1.39 (0.36) | 1.22 (0.31) |
| **LDL (mmol/L) [mean (SD)]** |  | 3.61 (0.84) | 3.65 (0.93) | 3.31 (0.96) |
| **TG/HDL ratio [mean (SD)]** |  | 1.29 (1.03) | 1.60 (1.16) | 2.18 (1.54) |
| **Systolic blood pressure (mmHg) [mean (SD)]** |  | 138.91 (19.58) | 143.59 (19.70) | 146.27 (19.59) |
| **Diastolic blood pressure (mmHg) [mean (SD)]** |  | 82.00 (10.65) | 83.05 (10.68) | 84.39 (11.01) |
| **CAD (%) [mean (SD)]** |  | 4561 (1.3) | 1031 (2.1) | 693 (6.2) |
| **CKD (%) [mean (SD)]** |  | 3902 (1.1) | 1113 (2.2) | 906 (7.5) |
| **NAFLD (%) [mean (SD)]** |  | 1119 (0.3) | 242 (0.5) | 407 (3.4) |

**Table 2.** Protein filtering across observational, interventional, and genetic evidence. Starting from Bonferroni-significant hallmark–protein associations in UK Biobank (row 1), proteins were successively filtered by concordance with semaglutide-induced changes in STEP 1/2 trials (row 2), nominal support from bidirectional Mendelian randomization (row 3), directional agreement between MR and observational estimates (row 4), and overlap with approved drug targets in DrugBank (row 5, excluding oncology agents; inhibitor + activator counts in parentheses). Columns stratify by hallmark and glycemic stage.

| Funnel stage | BMI \|<br>Normoglycemic | BMI \|<br>Prediabetes | BMI \| T2D | HbA1c \|<br>Normoglycemic | HbA1c \|<br>Prediabetes | HbA1c \| T2D |
| --- | --- | --- | --- | --- | --- | --- |
| 1. UKB reverse obs (Bonferroni < 0.05) | 2042 | 1265 | 526 | 1564 | 494 | 239 |
| 2. + STEP trial P < 0.05 | 657 | 482 | 240 | 401 | 164 | 102 |
| 3. With nominal reverse MR (IVW P < 0.05) | 176 | 146 | 90 | 8 | 1 | 2 |
| 4. Concordant: sign(rev MR IVW) = sign(UKB reverse obs) | 140 | 129 | 84 | 6 | 1 | 2 |
| 5. CAD DrugBank hits | 19 (19+0) | 19 (19+0) | 10 (10+0) | 2 (0+2) | 0 (0+0) | 2 (0+2) |

Protein associations showed varying degrees of consistency across stages (Fig 2B). TG/HDL and BMI had the largest number of directionally concordant associations shared across all three glycemic strata (792 and 620, respectively), while HbA1c associations were comparatively stage-specific, with a distinct set emerging only upon transition to T2D (Data S1).

**Figure 2.**
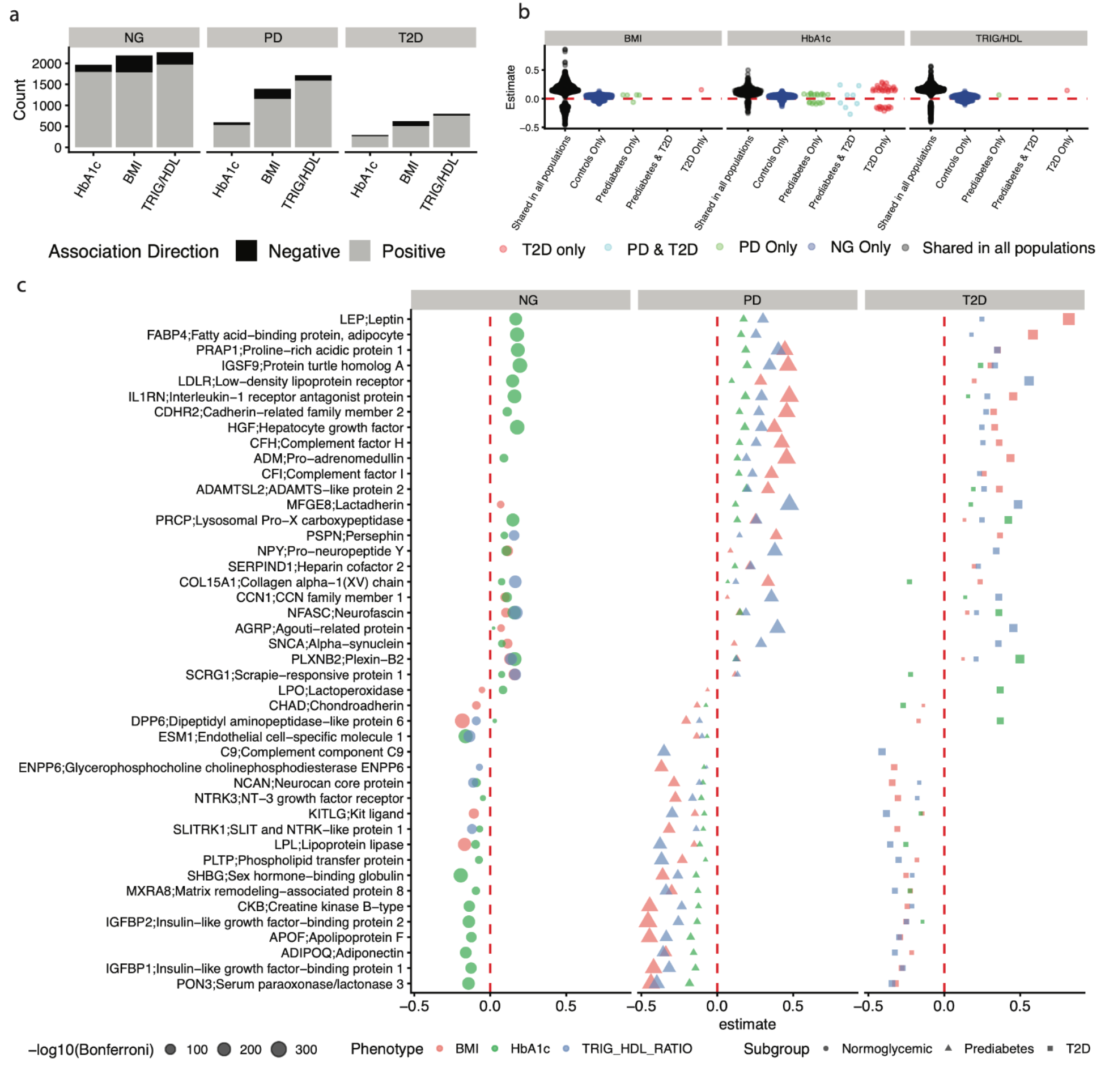
Proteomic associations with metabolic traits across glycemic stages. Proteins were associated with clinical indicators of metabolic dysfunction (e.g. BMI, HDL, HbA1c) across normoglycemic (Normo), prediabetes (PD), and type 2 diabetes (T2D) subgroups. (a) Number of significant protein–trait associations (FDR < 0.05) across glycemic stages, colored by direction of effect (positive, grey; negative, black). (b) Jitter plot showing protein–trait associations colored by subgroup specificity: T2D only (red), PD & T2D (light blue), PD only (green), Normo only (dark blue), and proteins shared across all subgroups (black). (c) Top protein–trait associations across glycemic stages. Point size reflects −log_10_(FDR-adjusted P); shape denotes glycemic subgroup; color denotes hallmark trait.

The largest effect sizes among Bonferroni-significant proteins were established metabolic regulators, including leptin (median absolute value of association, or |β| = 0.273), adiponectin (0.270), IGFBP1 (0.257), and growth hormone receptor (0.201) (Data S1) ^18,19–22^. The atlas also revealed associations with proteins less commonly linked to T2D in proteomic studies, including immune mediators (CD72, CD4, CD5), neurovascular signaling components (brain-specific angiogenesis inhibitor 1-associated protein 2, semaphorin-3F), and extracellular matrix proteins (ADAMTSL2). Sixteen proteins were associated with hallmarks only in prediabetes and T2D (e.g., interleukin-25, interleukin-3),highlighting potentially disease-specific pathways activated in stages beyond normoglycemia.

If the three hallmarks operated through identical proteomic pathways, their protein signatures would be redundant and a single axis would suffice. To test this, we computed pairwise Pearson correlations between vectors of protein-level effect estimates (betas) for each pair of hallmarks within each glycemic stage (Fig 3A). For example, within normoglycemia, we correlated the 2,923 protein betas for BMI against those for TG/HDL ratio, asking whether proteins strongly associated with one hallmark show similar associations with another.

**Figure 3.**
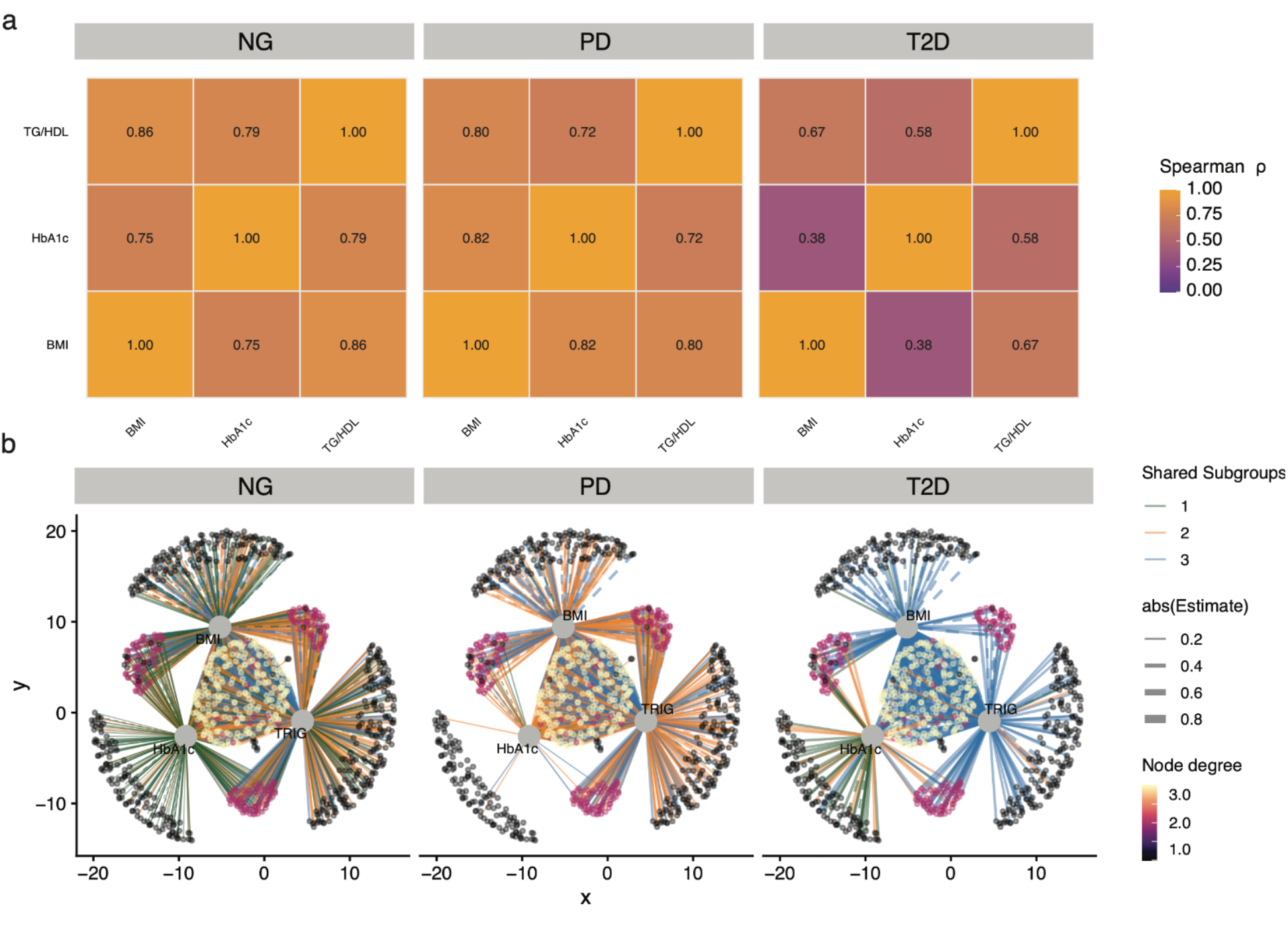
The architecture of metabolic disease across glycemic states. (a) Correlations between all metabolic hallmarks for each of the populations evaluated (i.e., people who had normoglycemia, prediabetes, or Type 2 Diabetes). (b) Network plot connecting proteins to metabolic traits, with edge thickness proportional to effect size. Edge color denotes glycemic status subgroup overlap (1: green, 2: orange, 3: blue).

Pairwise correlations between hallmark-specific protein effect estimates (Fig 3A) were highest between normoglycemia and prediabetes (ρ = 0.72-0.86 across BMI, HbA1c, and TG/HDL) and lowest in T2D, where the correlation between BMI- and HbA1c-aligned effects dropped to ρ = 0.38. This indicates that the cross-hallmark proteomic architecture is largely preserved across healthy and at-risk states (and hallmark) but reorganizes upon transition to overt T2D.

A protein-hallmark network (Fig 3B) revealed a core set of 21 proteins strongly associated across all hallmarks and stages with median |β| > 0.1 (exceeding the 73 percentile of all effect sizes; e.g., adiponectin, lipoprotein lipase, pro-adrenomedullin). HbA1c-specific associations emerged only in T2D (e.g., lactoperoxidase, membrane primary amine oxidase), while transitional associations shared between adjacent stages (e.g., angiopoietin-related protein 7, follistatin-related protein 1) tracked the proteomic stages toward T2D. Tissue-level analysis via GTEx integration (Supplemental Fig S1B) showed that most trait-associated proteins were abundant in liver, subcutaneous adipose, coronary artery, aorta, and lung, with a subset enriched in brain (e.g., reticulon-4 receptor, alpha-synuclein).

### Proteins that respond to GLP-1RA treatment and hallmark associations

To assess therapeutic response, we compared the “reverse” observational protein-hallmark associations from UKB with proteomic changes from the STEP 1 and STEP 2 semaglutide trials ^15^ (Fig 1B, Fig 4, see *Methods*). STEP 1 enrolled individuals with obesity but without T2D, while STEP 2 enrolled individuals with both T2D and obesity; both measured circulating proteomic changes at 68 weeks via SomaScan. In an RCT setting, one ascertains how GLP-1RA is associated with change in protein levels(e.g., GLP-1RA → protein). To make the observational analyses more comparable to this framework, we modeled protein levels as the outcome and each clinical hallmark as the exposure (e.g., BMI → protein [STEP1 comparison] or glycemic level → protein [STEP2 comparison]) [Data S1]. We selected the UKB glycemic stratum whose inclusion criteria parallel those of each trial: the normoglycemic and prediabetes strata for STEP 1 (individuals without T2D), and the T2D stratum for STEP 2 (individuals with established diabetes). These hallmark-first associations complement the protein-first analysis (Hallmark ∼ Protein, Figure 1A). This hallmark-first association is denoted by Protein ∼ Phenotype for three focal hallmarks, including BMI, HbA1c, and TG/HDL ratio, across all three glycemic strata. Whereas the protein-first beta quantifies the difference in a hallmark per standard deviation of protein, the reverse beta represents the difference in protein level per standard deviation of the hallmark, providing a common scale for integration with semaglutide trial effects and genetic estimates below. Both are cross-sectional associations estimated across participants at a single timepoint.

**Figure 4.**
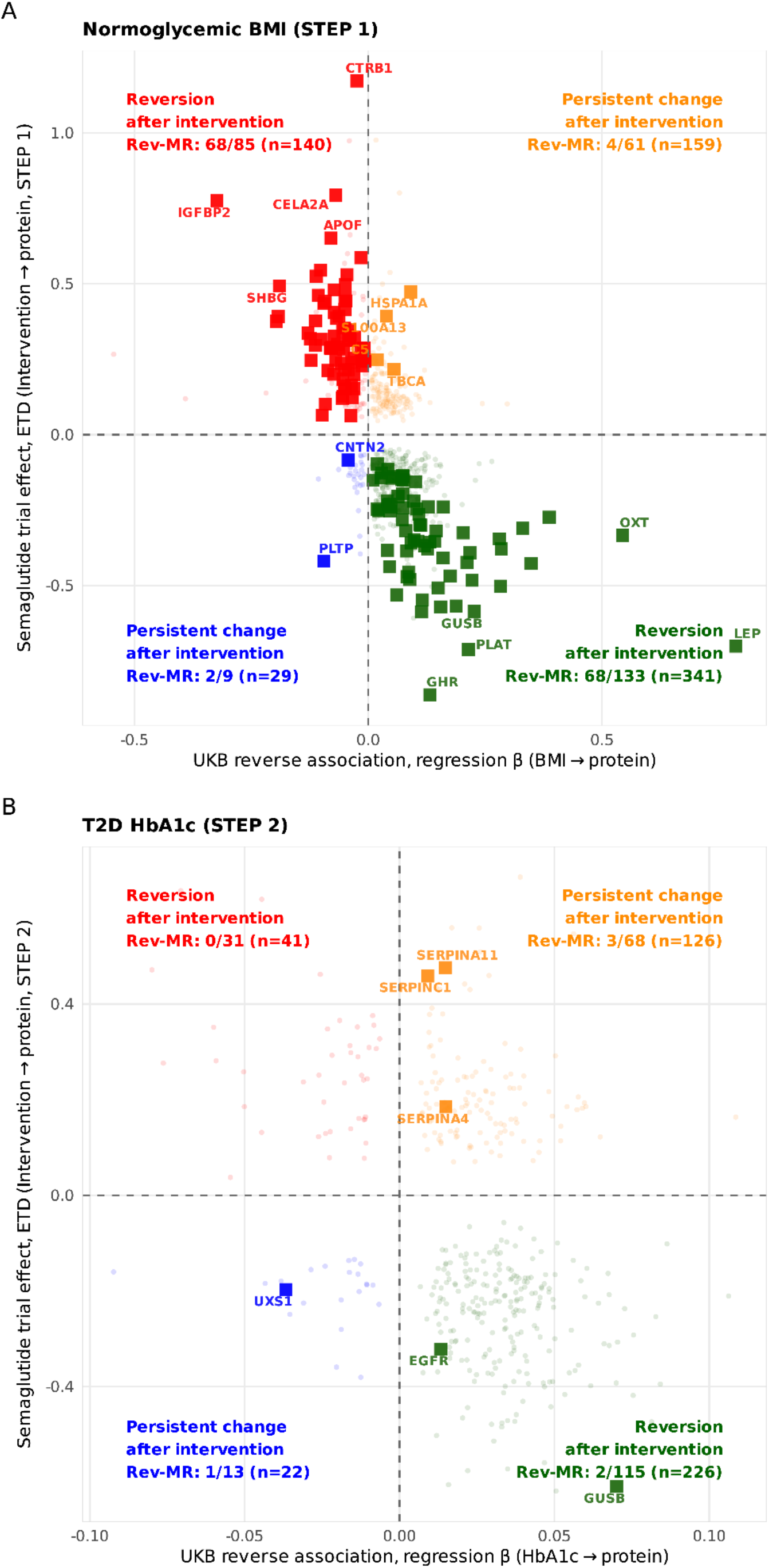
Proteins reversed or persistent to GLP-1 receptor agonist treatment. (a–b) Scatterplots of HbA1c-associated proteins in UK Biobank versus post-treatment change in STEP 1 (inclusion criteria were individuals without T2D but with obesity) (a) and STEP 2 (inclusion criteria was T2D) (b). Quadrants classify proteins by concordant or discordant directionality: reversion (red, green) or persistence (orange, blue). Red denotes UKB-/GLP-1RA+; green denotes UKB+/GLP-1RA-; yellow: UKB+/GLP-1RA+; blue: UKB-;GLP-1RA-. Square indicates a significant hallmark-first (hallmark → protein) MR association **concordant** with direction of the UKB and STEP trial. ETD: Estimated Treatment Difference.

Among 2,923 circulating proteins measured by Olink in UK Biobank (of which 2,142 had gene-symbol matches to the SomaScan platform used in STEP 1/2), 960 (32.8%) showed stringent hallmark → protein association for BMI, HbA1c, or triglyceride/HDL cholesterol and nominal (p-value < 0.05) semaglutide-associated change in at least one trial (STEP 1 or STEP 2) (Table 2). In normoglycaemia, TG/HDL and BMI contributed the largest trial-overlap layers (859 and 657 proteins with nominal semaglutide p-value < 0.05 on the hallmark-aligned trial contrast, respectively), compared with HbA1c (401) Counts declined at higher glycaemic stages (Table 2).

Proteome-wide associations between BMI-, TG/HDL, or HbA1c-protein estimates in UKB and semaglutide-protein effects were strongly inversely correlated (Pearson r ranging from −0.56 to −0.36, both light and dark points), confirming that proteins that are correlated with hallmarks in an observational or cross-sectional setting (e.g., BMI or glycemic trait) are broadly concordant with therapy-induced response (Fig 4A-B). We mapped proteins into four quadrants based on joint directionality in UKB and STEP, distinguishing "reversion" proteins (whose hallmark-associated direction was reversed by semaglutide, suggesting therapeutic responsiveness) from "persistent" proteins (whose direction was maintained under treatment, suggesting dysregulation not normalized by GLP-1RA). Among individuals with normoglycemia, 669 proteins were modulated by semaglutide in STEP 1: 481 fell into hypothetical “reversion” quadrants (e.g., GHR, LEP) while 188 were “persistent” (e.g., HSPA1A, S100A13). In the T2D subgroup, 415 proteins were modulated in STEP 2: 267 reverted while 148 remained persistent (e.g., EGFR, SERPINA11).

Proteins that change under GLP-1RA intervention may reflect direct drug effects or downstream consequences of improved hallmark levels (Figure 1B). To disentangle these possibilities and identify proteins with genetic evidence for a causal relationship with hallmark variation, we performed bidirectional Mendelian randomization (MR) (Figure 1B,C). MR uses genetic variants as instruments to test whether genetically predicted protein levels causally affect hallmark traits (protein-first direction) or whether genetically predicted hallmark variation causally alters protein levels (hallmark-first direction), independent of drug exposure. We performed MR using a split-sample design: cis-pQTL instruments from the UKB Olink subsample (∼55,000) with phenotype GWAS in the held-out sample (∼418,000), ensuring no sample overlap (see Methods, Fig 1C). We tested all 2,923 proteins for which at least one genome-wide significant (p < 5×10^-8^) cis-pQTL instrument was available.

We conducted genome-wide association studies (GWASs) of metabolic hallmarks conducted in approximately 418,000 UK Biobank participants who did not overlap with the Olink subsample. GWASs identified 46,081, 57,512, and 35,951 genome-wide significant (p-value < 5 x 10^-8^) for BMI, HbA1c, and the TG/HDL ratio, respectively (Supplemental Figure S2). To identify quasi-independent signals, genome-wide significant variants were ranked by association strength. Starting with the most significant variant, we retained that variant as a sentinel signal and excluded any additional genome-wide significant variants located within 500 kb on the same chromosome, yielding 612, 2,858, and 1,664 for BMI, HbA1c, and TG/HDL, respectively (Methods). The strongest associations localized to well-known metabolic regions: rs1421085 at the *FTO* locus confirming previously association with BMI ^23^, rs17476364 lead variant in HK1 locus for HbA1c, previously known to be associated with reduced T2D risk and HbA1c ^24,25^, and rs964184 at the hepatic 11q23 APOA1/APOC3/APOA5 cluster for TG/HDL. Together, these GWAS summary statistics supplied the held-out, trait-specific GWAS for bidirectional MR against cis-pQTL instruments generated from Olink proteomics subset.

Table 2, rows 3 and 4, display the number of proteins identified by the MR process that were also triangulated by the RCTs. In total, we found that most “concordant” proteomic shifts, e.g., proteins (62.4% [136 out of 218 MR-testable proteins])) in the two reversion quadrants of Fig 4a-b (red and green), that travel upward with BMI (or HbA1c %) and downward with GLP-1RA, possessed genetic evidence. We revealed three distinct genetically-derived architectures (Supplemental Figures S3-S5). For BMI, “reverse causative” dominated: 106 proteins were causally altered by adiposity, while only 10 showed protein-first effects (protein → BMI). For HbA1c, the pattern inverted: 19 proteins causally influenced glycemia in the protein-first direction, but zero were altered by HbA1c in the reverse direction. TG/HDL ratio showed a mixed architecture: 17 protein-first and 34 hallmark-first significant proteins, with 5 showing concordant bidirectional effects (both protein → HbA1c and HbA1c → protein) (e.g., PLTP, SHBG).

We next compared the direction of MR estimates with STEP trial data (Fig 4A-B). Because triple triangulation requires protein measurements on both the Olink (UKB) and SomaScan (STEP, deCODE) platforms, it was possible only for the 2,142 cross-platform proteins; proteins measured exclusively on one platform could not be assessed for convergence across all three evidence streams, and their absence from the triple-triangulated set should not be interpreted as failure to triangulate. For BMI, 142 of 669 significant proteins showed concordance across all three evidence streams. We defined as directionally consistent and at least nominally significant (p < 0.05) estimates in each layer: observational association (UK Biobank [BMI → protein]), MR causal estimate (for BMI change → protein), and experimental *reversal* under semaglutide. “Triple-triangulated” proteins (e.g., those in Table 2, row 3-4) for BMI change inducing protein change included leptin (observational beta = 0.79, MR beta = 0.0457, STEP 1 effect = −0.70), IGFBP2, (observational beta = −0.323, MR beta = −0.023, STEP 1 effect = 0.775) and GHR (observational beta = 0.0304, MR beta = 0.0199, STEP 1 effect = −0.862). For example, in the STEP1 trial, GLP-1RA causes the protein leptin, a known causal factor for obesity ^26^, to *decrease* its levels in the blood (Fig 4a). In the UK Biobank, an increase in BMI is genetically causally related to increased levels of Leptin (LEP, x-axis, Fig 4a, solid square point). Analogous interpretations can be made for the other proteins in Fig 4a.

For HbA1c %, 2 proteins were triple-triangulated (Table 2, row 4), or found in the observational UKB, semaglutide RCT (STEP2) (Figure 1A, B, and C). These proteins included EGFR and GUSB (Figure 4B). For TG/HDL ratio, 57 concordant proteins were double triangulated (Table 2, rows 1 and 2), including CPB1, SHBG, and adiponectin (Data S1, Supplemental Figure S5).

### Persistent pathways are predictive of cardiovascular disease

We noted that many of the pathways represented by the persistent proteins (e.g., those whose direction of association in UKB MAP-D is not reversed by semaglutide in STEP1/2) could index pathways that are associated with T2D macrovascular complications, namely heart disease. To test this, we computed associations between these trial-triangulated proteins (Figure 1D) and incident coronary artery disease (CAD; new diagnoses occurring after the baseline assessment, excluding individuals with prevalent CAD at or before baseline) in the UKB Olink proteomics subsample (n ≈ 44,000 with complete covariates) (Figure 5A), using logistic regression that adjusted for age, sex, glycemic stratum, assessment center, BMI, HbA1c, and 40 genetic PCs (see Methods).

**Figure 5.**
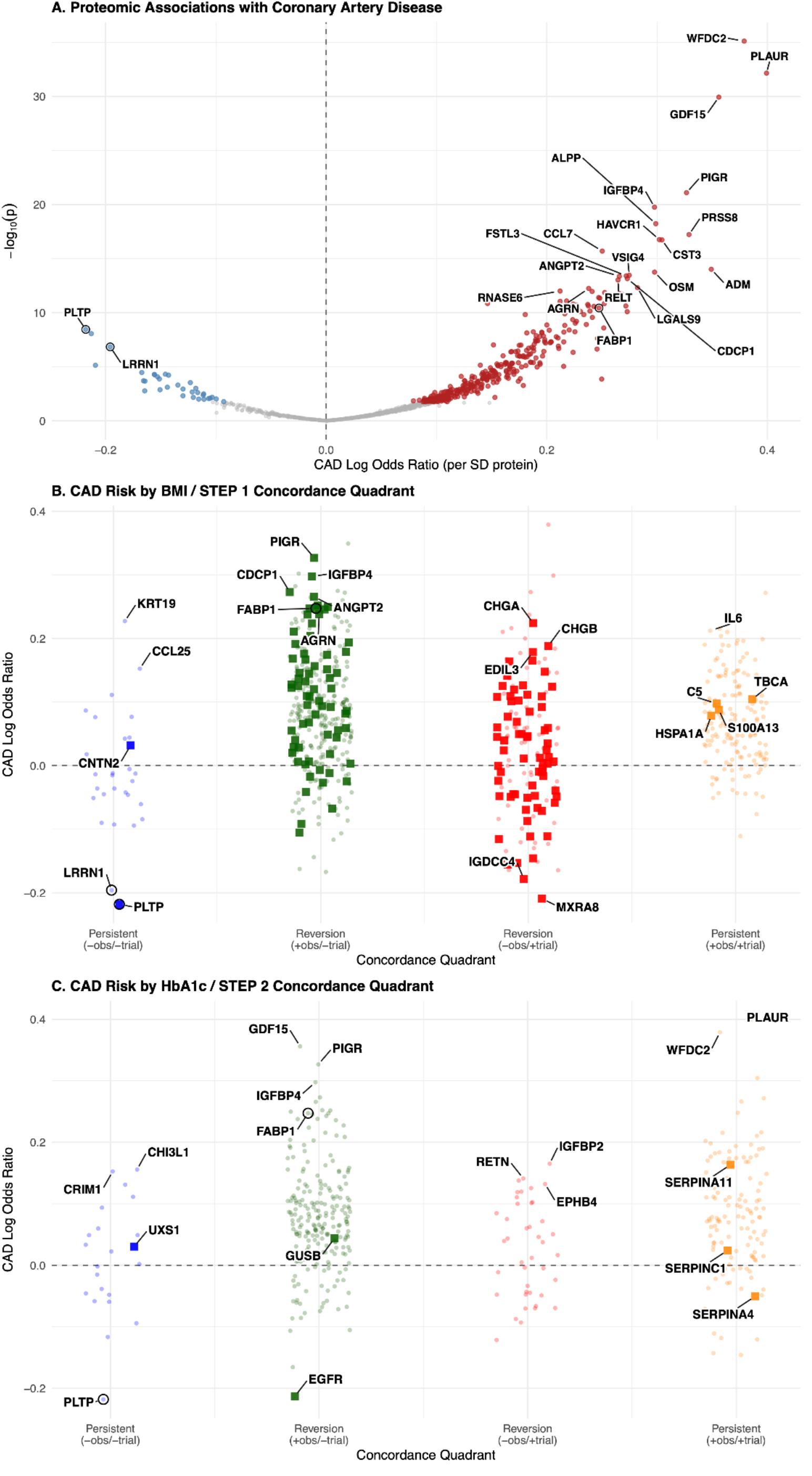
GLP-1RA–persistent proteins are associated with coronary artery disease. (a) Protein associations with incident coronary artery disease (CAD) among validated proteins. Point size reflects −log_10_(Bonferroni-adjusted p-value); color and axis denote log_10_ odds ratios. (b) Distribution of complication associations stratified by quadrants displayed in Figure 4 for Step1 validated proteins. X axes are coefficients from the volcano plot in the same row. Y-axes correspond to the quadrant in which a given protein appeared in Figure 4. Small black points are insignificant associations. Large points were FDR-significant. (c) same as (b), but for Step 2 validated proteins and quadrants. CAD: Coronary Artery Disease

Persistent proteins were enriched for incident CAD risk signals. Stratifying the 1,008 proteins by their Fig 4 quadrant (Fig 5B-5C), all proteins in the upper right "persistent" (+UKB/+trial) quadrant showed a positive point risk estimate for incident CAD (e.g., IL6, HSPA1A, S1000A13, C5, TBCA for STEP 1; PLAUR, WFDC2, SERPINA11 for STEP2). Among these persistent GLP-1RA-non-responsive proteins, the top individual associations with CAD included SCARB2 (OR of 1.2, FDR-corrected p-value < 1×10^-6^) and TAFA5 (OR of 1.2, FDR < 1×10^-6^).

Conversely, proteins that were inversely associated with BMI in UKB and remained suppressed under semaglutide in STEP1 (e.g., the persistent (-UKB/-trial) quadrant of Fig 5B) were associated with increased CAD risk, meaning that lower circulating levels corresponded to higher disease odds. PLTP (OR: 0.804, FDR corrected p-value: 6.7×10^-8^) and LRRN1 (OR: 0.822, FDR corrected p-value: 1.72×10^-6^) were among the strongest inverse associations for CAD.

We further evaluated whether the proteins significantly associated with incident cardiovascular disease could be matched to pharmacologic agents with directionally consistent actions using DrugBank-derived annotations. For proteins with positive associations (i.e. higher protein levels associated with higher disease odds), we prioritized inhibitors or antagonists. Conversely, for proteins with inverse associations (i.e., lower protein levels associated with higher disease odds), we prioritized agonists or activators. For example, FABP1, positively associated with CAD risk (OR = 1.28), was matched to two approved inhibitors, including oleic acid and fenofibric acid which *reduce* the protein’s activity^27^. HNMT, a histamine-metabolizing enzyme (OR = 1.14), was matched to amodiaquine and diphenhydramine. HNMT has limited prior evidence linking it to coronary artery disease, suggesting the association may represent underexplored therapeutic axes for residual cardiovascular risk, such as inflammation. A complete list of matched pharmacologic candidates is provided in Data S1.

## Discussion

The stages that are defined by normoglycemia to prediabetes to diabetes is commonly defined by glucose and glycemic control ^28^; yet upstream risk is influenced by multiple metabolic processes, including adiposity, insulin resistance, and glycemia^4,29^. We found that these cardiometabolic hallmarks have distinct causal relationships with the circulating proteome. Adiposity predominantly reshapes protein levels (hallmark-first causative), consistent with adipose tissue’s established role as an endocrine organ that broadly remodels circulating protein profiles. By contrast, the protein-first architecture observed for glycemia raises the possibility that circulating proteins provide mechanistic insight into biological pathways driving dysglycemia, rather than simply reflecting glucose exposure - a distinction with implications for biomarker interpretation and therapeutic target selection. Insulin resistance, as indexed by TG/HDL ratio, exhibited evidence of bidirectional feedback. That not all hallmark-associated proteins normalize under semaglutide further suggests that some biological pathways are incompletely addressed by GLP-1RA monotherapy, potentially reflecting residual features of the obesity or insulin resistance phenotype that persist despite weight loss. These findings highlight the potential of proteomic atlases to capture disease heterogeneity beyond glycemic status ^30,31^. We arrived at these conclusions through *triangulation* combining evidence from cross-sectional associational data, randomized trials, and genetic associations to prioritize proteins and pathways across the vast atlas of possible relationships ^17,32^.

T2D emerges through heterogeneous trajectories that are only partly reflected by glycemic labels ^1,4,29,33^ and genome-wide association study implicated pathways ^2^. MAP-D addresses this by defining circulating protein-hallmark relationships across normoglycemia, prediabetes, and incident diabetes, yielding a circulating proteomic view of how metabolic biology varies with glycemic stage. Importantly, when paired with GLP-1RA trial contrast ^15^, it becomes possible to classify proteins according to whether hallmark-associated patterns can be potentially reversed by pharmacologic exposure or are persistent, a distinction that existing biobank-scale proteomic studies cannot make.

Split-sample Mendelian randomization^34,35^ adds complementary information about bidirectional architectures linking proteins and hallmarks, helping separate patterns dominated by adiposity-driven proteomic remodeling from those in which upstream protein variation drives glycemic control, with insulin resistance-related pathways (e.g., TG/HDL ratio) exhibiting bidirectional feedback. In combination, stage-stratified association mapping, intervention-responsive protein shifts, and genetic directionality provide complementary constraints on interpretation. We believe that observational structure places proteins in metabolic context; trial data tests sensitivity to GLP-1RA; genetics probes stable, instrument-supported directional relationships^32^. This integration is particularly suited to disambiguating drug-responsive biology from correlates of glycemic stages.

A central implication of the reversion-persistence framing is that proteomic “alignment” with BMI, HbA1c, or TG/HDL ratio does not necessarily collapse onto a single interpretive axis once GLP-1RA induced proteomic remodeling or genetic-based causative associations is considered. Proteins that remain discordant with therapeutic direction may reflect remodeling, inflammatory or stress programs ^36^ leftover from getting to the obesity or glycemic state, organ-specific regulation, or treatment-independent biological states that are imperfectly indexed by cross-sectional hallmark levels and/or precisely the class of biology that might contribute to residual risk ^37^, if it is not “normalized” by interventions ^38^ ^39^.

Persistent proteins may be patient-specific biomarkers for both the efficacy of a drug in treating phenotypic hallmarks (e.g., obesity) while also classifying patients as potentially at-risk for complications. Certain persistent proteins are targets of approved treatments, such as EPHB4 ^27^. These findings suggest that pharmacologic modulation of intervention-persistent proteins may complement single agonist GLP-1RA-based interventions, either by mitigating residual risk or by enhancing efficacy in partial responders. Clinical trials, such as SELECT^40^, FLOW ^41^ and SUSTAIN-6 in type 2 diabetes ^42^, demonstrate population-level cardiovascular and renal disease risk reductions of 20–26%. However, primary events persist in both treatment and placebo arms, underscoring the need to investigate prior history and incomplete therapeutic responses. Head-to-head evidence that reduction beyond what is observed with single agonist GLP1-based interventions is a strong case for the possibility for improvement, such as data emerging from the *multiple-agonist* GLP1 drugs such as tirzepatide ^43^. Several of the persistent proteins most strongly associated with incident CAD index inflammatory and cellular-stress programs rather than hallmark severity per se. GDF15, a stress-responsive cytokine, and IL6, a canonical mediator of the inflammatory contribution to atherosclerosis. Their dysregulation is not “reversed” by semaglutide in our analysis, consistent with the persistence of an inflammatory residual-risk axis that glucose- and weight-lowering does not fully normalize ^37,38^.

We found that most of the evidence points to adiposity associated with changes in protein levels; in contrast, we found that changes in protein levels are associated with changes in hemoglobin A1C. Among candidates prioritized by converging observational and pharmacologic evidence, GUSB (β-glucuronidase) and EGFR stand out. Both proteins were positively associated with HbA1c in UK Biobank and were lowered by semaglutide (Figure 1 A,B), with trial effects that were larger in individuals with *established* T2D (STEP 2) than in participants with obesity and normoglycemia (STEP 1). This suggests that their circulating levels track specifically with glycemic pathology rather than adiposity alone. GUSB is a lysosomal hydrolase released into circulation during lysosomal exocytosis and cellular stress; elevated plasma levels have been documented in diabetes ^44^. Increased GUSB in T2D ^45^ is consistent with chronic lysosomal and autophagic stress in β-cells, endothelium, and glomerular cells, and its suppression under semaglutide may reflect restoration of reduction of hyperglycemia-driven lysosomal damage. That GUSB additionally carries a directionally concordant protein-first Mendelian randomization signal strengthens the case for a causal role in glycemic control (Figure 1 C,D), whereas EGFR’s null protein-first MR is consistent with its circulating abundance being a downstream marker of inflammatory and metabolic state (a biomarker) rather than a driver of HbA1c. The pair shows a useful asymmetry: proteins like GUSB nominate potential causal nodes for further instrumented study (e.g., tissue-specific genetic perturbation of glycosaminoglycan catabolism), while proteins like EGFR are better interpreted as robust pharmacodynamic biomarkers.

TG/HDL was distinguished by a bidirectional architecture, exemplified by SHBG. SHBG carried concordant protein-first and hallmark-first genetic signals in our analysis, consistent with a feedback relationship between insulin resistance and the proteome rather than a simple cause-or-consequence relationship. Genetically higher SHBG lowers T2D risk ^46,47^ and more recent work suggests that it is mediated by visceral and liver fat ^48^.

### Limitations of the study

Much of the evidence behind MAP-D (UKB) and GLP-1RA-associated proteins, while directionally “concordant,” was not supported by genetic variants that linked protein to hallmarks. We view this as a complementary discovery layer. GWAS of GLP-1RA response has so far explained only a modest fraction of inter-individual variance in weight and glycemic outcomes, and germline pQTL maps are necessarily silent on biology revealed only upon drug exposure, at specific tissue sites, or in transient regulatory states. Concordant-but-not-MR-supported proteins in MAP-D (Figure 2, and Figure 4, light points) therefore nominate a complementary set of candidates those whose relevance may depend on induction by GLP1R agonism, cell-type-specific expression, environmental exposures, or post-translational regulation that lifetime genetic variation cannot instrument. Several parallel explanations remain worth testing: residual confounding from shared upstream regulators of adiposity and insulin resistance, concurrent medications, or lifestyle exposures. Of course, there is imperfect mapping of circulating abundance to tissue-specific activity in islet β-cells, hypothalamic neurons, enteroendocrine L-cells, adipocytes, and hepatocytes where GLP1R signaling is enacted.

Our focus on incident T2D enabled a clearer view of proteomic changes preceding disease onset, minimizing confounding from pharmacologic treatment and reverse causality. However, this may limit direct comparability to intervention trials such as STEP 2, which enrolled individuals with established T2D. Olink proximity extension assays and SomaScan aptamer-based assays quantify overlapping but not identical protein sets (2,142 of 2,923 UKB proteins had gene-symbol matches to SomaScan), and cross-platform discordance may inflate or obscure true concordance rates^49,50–52^. Observational proteomic associations and randomized treatment effects estimate related but distinct quantities; concordance across layers supports biological coherence rather than numeric interchangeability^32^. Mendelian randomization mitigates some confounding but remains sensitive to instrument validity, pleiotropy, and contextual stability of genetic effects^53,54^. Together, we consider MAP-D as an integrative resource most appropriately extended through experimental follow-up and prospective outcome studies.

In summary, we have built a comprehensive proteomic map of the hallmarks of T2D, leveraging biobanked proteomics and genetics. We showed how this resource could potentially be used to discriminate patient response to treatment while also providing additional therapies to minimize complications and optimize reversion to a healthy metabolic state. Future studies aim to expand the cohorts and hallmarks (e.g, HOMA-IR) analyzed in the MAP-D while integrating additional data types (e.g., metabolomics, microbiome) from diabetes-relevant tissues.

## RESOURCE AVAILABILITY

### Lead contact

Further information and requests for resources should be directed to and will be fulfilled by the lead contacts, Braden T Tierney and Chirag J Patel.

### Materials availability

This study did not generate new unique reagents.

### Data and code availability

The data from the UK Biobank that support the findings of this study are available upon application (https://www.ukbiobank.ac.uk/register-apply/). Code for this manuscript is available via Zenodo at https://doi.org/10.5281/zenodo.1707108755 and https://github.com/stejat98/MAP-D. The MAP-D atlas data are available at https://doi.org/10.6084/m9.figshare.30007306 ^56^. Additionally we make the GWAS summary statistics for the BMI, HbA1c, and TG/HDL Ratio hallmarks available at: https://doi.org/10.6084/m9.figshare.32048550 ^57^. An interactive app can be found here: https://mapd.bio and https://chiragjp-mapd.share.connect.posit.cloud/.

## ACKNOWLEDGEMENTS

C.J.P., S.I., F.B., S.T., J.M., B.T.T. received funding from the NIH National Institutes of Diabetes and Digestive, and Kidney Diseases (R01DK137993). C.J.P., S.I., S.T., G.W.M., B.T.T. received funding from Advanced Research Projects Agency for Health (ARPA-H) D24AC00345. C.J.P and S.T. received funding from the NIH National Institute of Environment Health Sciences (R01ES032470).

## Author contributions

B.T.T. and C.J.P. conceptualized and supervised the study and led the writing - original draft, review & editing. S.T. led the analysis, software development, visualization, and writing - original draft. S.I., Y.G., F.R., and P.G. contributed to formal analysis and writing - review & editing. A.L., J.M., and M.S.G. contributed to writing - review & editing. F.B. contributed to writing - review & editing. G.W.M. and V.T. contributed to writing - review & editing.

## Declaration of interests

The authors declare no competing interests.

## Supplemental information

Document S1. Supplemental Figures S1-S2

Data S1. MAP-D Atlas data and comparison with genetic evidence and STEP1/2 trials.

## STAR Methods

### EXPERIMENTAL MODEL AND STUDY PARTICIPANT DETAILS

#### Study sample

This study was conducted using data from the UK Biobank, a prospective cohort of approximately 500,000 participants aged 40-69 years at recruitment. Our primary analytic sample consisted of 474,318 White European participants, as defined by self-reported ancestry ("British," "Irish," "Any other White background"). Within this population, 12,068 individuals developed incident type 2 diabetes (T2D) over the follow-up period, identified through electronic health records, self-report, hospital admissions data, and primary care records. Proteomic data were available for ∼50,000 participants, of whom ∼42,000 White European participants had both proteomic and relevant cardiometabolic phenotypic data available for analysis.

#### Metabolic hallmarks in the UK Biobank

We assessed three key cardiometabolic hallmarks of diabetes at the first assessment visit along the axes of adiposity, glycemic control, and insulin resistance: body mass index (BMI) (kg/m^2^), hemoglobin A1C% triglyceride-to-HDL cholesterol ratio (TG/HDL ratio).

#### Metabolic stage in the UK Biobank

In the UK Biobank (Figure 1A), we defined glycemic status into three mutually exclusive stratum: normoglycemia, prediabetes, and incident type 2 diabetes (T2D). Participants with diagnosed ICD10 codes (E110, E112, E113, E114, E115, E116, E117, E118, and E119) or self-reported diabetes (UK Biobank self-reported code: 1223) at baseline, were excluded to avoid confounding from treatment or disease duration. Incident T2D was defined as a new diagnosis occurring during follow-up, enabling modeling of proteomic changes that precede disease onset. Normoglycemia was defined as HbA1c <39 mmol/mol (<5.7%). Prediabetes was defined as HbA1c 39–48 mmol/mol (5.7-6.5%) without a clinical diagnosis of diabetes or diabetes medication. Incident T2D was defined as a new ICD-10 diabetes diagnosis during follow-up or HbA1c >48 mmol/mol (>6.5%); individuals with an ICD-10 diabetes diagnosis but HbA1c ≤ 48 mmol/mol who lacked diabetes medication were classified as prediabetes.

#### UK Biobank Proteomics

Proteomic profiling was performed using the OLINK Explore 3072 platform, a high-throughput proximity extension assay (PEA) technology that quantifies 2,923 circulating proteins ^58^. Protein levels were measured in plasma samples collected at UK Biobank assessment centers, with quality control performed by OLINK to ensure consistency and accuracy. Protein expression levels were log2-transformed and normalized by OLINK to account for batch effects before being made available for analysis.

#### STEP 1/2 Randomized Clinical Trial and Proteomics

STEP1 and STEP2^15,59^ were randomized controlled trials aimed to measure the efficacy of GLP-1RA interventions to lower cardiometabolic risk (e.g., glycemic levels, weight, and risk for cardiometabolic disease) on individuals with obesity and without T2D (STEP1) and individuals with both T2D and with obesity (versus standard of care, STEP2). STEP 1 included 1,311 participants with a mean age (years) of 47.5 (standard deviation [SD]: 12.7), 72.8% (n=955) females, a mean BMI (kg/m^2^) of 37.9 (SD: 6.7) and a mean HbA1c (%) 5.7 (SD: 0.3). STEP 2 included 645 participants with a mean age (years) of 56.3 (SD: 10.8), 49.8% (n=321) females, a mean BMI (kg/m^2^) of 35.7 (SD: 6.5) and a mean HbA1c (%) 8.1 (SD: 0.8). Plasma proteomics in STEP 1 (n = 1,311 participants with obesity without T2D) and STEP 2 (n = 645 participants with T2D) were profiled using the SomaScan v4.1 aptamer-based platform (SomaLogic), which measures approximately 7,000 protein analytes (6,386 unique protein targets). Semaglutide-induced protein changes (log_2_ fold-change, treatment vs. placebo at 68 weeks) were used as the experimental comparator. Of the 2,923 UKB Olink proteins and 6,386 unique protein targets, 2,142 proteins were mapped across platforms by gene symbol and used for UKB-STEP comparisons.

#### External plasma pQTL summary statistics (deCODE)

For reverse Mendelian randomization (trait → protein), we required protein-level genetic association summary statistics from a cohort independent of UK Biobank. We used plasma pQTL results from the deCODE genetics study ^31^, which measured 4,907 aptamers in 35,559 Icelandic individuals using the SomaScan aptamer-based proteomics platform. These summary statistics provide genome-wide variant-protein associations suitable as outcome data in two-sample MR, where UK Biobank phenotype GWAS instruments (derived from held-out non-proteomics sample) serve as exposures.

### METHOD DETAILS

#### Mapping the proteome across hallmarks within glycemic stratum in the UK Biobank

We describe each stage of derivation of MAP-D and triangulation approaches (Figure 1A-C). First, a proteome-wide association study (PWAS) was conducted to systematically evaluate associations between each of the 2,923 proteogenomic indicators of protein levels and the three cardiometabolic hallmarks (BMI, TG/HDL ratio, hemoglobin A1C). All statistical analyses were performed in R (version 3.6.1). Linear regression models were applied to estimate the association between each protein and each cardiometabolic outcome. Outcome and protein variables were standardized using the scale() function in R, which centers the variable by subtracting the mean and scales it by dividing by the standard deviation. Models were adjusted for key covariates including age (years), sex (male/female), assessment center (to account for geographical differences), household income (as a proxy for socioeconomic status), fasting time, and 40 genetic principal components.

For each of 2,923 circulating protein analytes, we estimated two complementary linear associations with each of the three cardiometabolic hallmark traits, analyzed separately within three glycemic stages (normoglycemia, prediabetes, and type 2 diabetes). First, we computed “protein-first” associations with each hallmark as the outcome and each protein as the exposure: Hallmark ∼ I(scale(protein)) + covariates. Second, we computed “hallmark-first” associations with each protein as the outcome and each hallmark as the exposure: Protein ∼ I(scale(hallmark)) + covariates.

Multiple testing correction was applied using the Bonferroni correction, setting the significance threshold at p-value < 0.05 / (26,307) to account for the large number of protein-trait associations tested across 3 stages and 3 hallmark indicators.

#### Integration with STEP 1 and STEP 2 Trial Proteomics

We compared UKB-derived protein associations with those observed in the STEP 1 (obesity and non-T2D) and STEP 2 (T2D) semaglutide trials^15,59^ (Figure 1B). Among the 2,142 proteins that were mapped across platforms by gene symbol and used for UKB-STEP comparisons, 1008 unique proteins were nominally significant in at least one trial (STEP 1 or STEP 2) and used for all STEP 1/2 comparisons and downstream analyses. Proteins were grouped into: concordant in direction and significance, concordant in direction only, and discordant in direction. We also assessed which proteins were reversed by semaglutide in direction compared to their UKB association, as a proxy for therapeutic responsiveness.

#### Pathway Enrichment Analysis

Gene identifiers for bonferroni-significant MAP-D proteins (across all hallmarks) were subjected to GO Term enrichment analysis (using the 2023 GO Biological Process Database) with the enrichr R package with the default settings^60^. Enrichment was performed separately for the groups with normoglycemia, prediabetes, and T2D to identify shared and divergent biological mechanisms.

To evaluate the tissues linked to circulating proteins associated with metabolic stage, we leveraged GTEx, a resource profiling gene expression across 54 tissues from 838 donors with matched genotype data. Using GTEx-defined expression specificity, we assigned proteins to a given tissue if their corresponding gene exhibited at least fourfold higher expression in that tissue compared to the mean expression across all others, consistent with prior strategies for mapping plasma proteins to organ-level sources ^61^. This enabled us to classify validated proteins into tissue-specific sets while reducing bias from organs represented by multiple sub-tissues (e.g., brain, which is divided into several cortical and subcortical regions). We then tested for enrichment of these tissue-specific protein sets among MAP-D-validated proteins, providing insight into the organ systems most implicated in stages of metabolic disease and therapeutic intransigence.

#### Genome-wide association studies for split-sample Mendelian randomization

We conducted genome-wide association studies (GWASs) using REGENIE v4.1.2 ^35^. Protein GWAS were performed in the Olink proteomics subsample (n ∼ 55,000), and hallmark phenotype GWAS were performed in the held-out sample *without* proteomics (n ∼ 418,000), ensuring no sample overlap. REGENIE step 1 fitted null models using non-imputed genotypes with block size 1000, and step 2 performed association testing on imputed genotypes (∼1.2M variants per chromosome) with block size 400. Covariates included age, sex, 40 genetic PCs, assessment center, and household income.

We performed bidirectional Mendelian randomization ^34^ to test if proteins were genetically causing changes in hallmarks, or if hallmarks caused changes in protein levels. Bidirectional MR here denotes testing both pre-specified directions of effect; the designs are two-sample throughout, with protein-first MR using non-overlapping UK Biobank subsets and hallmark-first MR using UK Biobank trait instruments with external protein quantitative trait summary statistics to reduce bias of overlapping sample ^62^. For protein-first MR (protein in association with phenotype), cis-pQTLs were defined as genome-wide significant (p-value < 5 x 10^-8^) variants within 500 kb of the encoding gene, clumped at r^2^ < 0.001 using 1000 Genomes European reference. The Wald ratio was used for proteins with a single instrument; inverse-variance weighted (IVW) MR was used for proteins with greater than or equal to 2 instruments ^63^. For hallmark-first MR (hallmark in association with protein), we used genome-wide significant SNPs (p-value < 5 x 10^-8^) from the same held-out phenotype GWAS as candidate instruments, retaining quasi-independent signals by iteratively keeping the strongest variant and excluding additional variants within 500 kb on the same chromosome. These trait instruments were harmonized to protein outcome summary data from external DECODE SomaScan cis-pQTL results (Iceland) ^31^. After defining quasi-independent genome-wide significant trait instruments in the UK Biobank (612, 2,858, and 1,664 for BMI, HbA1c, and TG/HDL, respectively, under 500 kb pruning), 422, 282, and 259 SNPs remained for the primary hallmark-first-MR IVW after restricting to SNPs available in the external protein outcome summaries and harmonizing alleles. IVW MR was the primary method; MR-Egger was used as sensitivity analysis to detect directional pleiotropy ^64^. Cochran’s Q statistic assessed heterogeneity.

#### Association of RCT-triangulated proteins with Incident CAD

To evaluate whether the STEP trial-triangulated proteins were individually associated with cardiovascular risk, we tested each of 1,008 (out of 2,142 overlapping measured proteins) triangulated proteins - defined as those significantly associated with at least one metabolic hallmark in MAP-D and nominally modulated by semaglutide in STEP 1 or STEP 2 (p-value < 0.05) across all glycemic strata - for association with incident coronary artery disease (CAD) using logistic regression in the UK Biobank Olink proteomics resource (i.e. participants with usable proteomics data in the analysis dataset), not the full UK Biobank cohort. We excluded individuals with prevalent CAD at or before the baseline assessment; incident CAD cases were participants who developed CAD after baseline under the ascertainment rules below, and controls were participants without incident CAD through follow-up among those retained. Models were fit per protein using complete cases for binary CAD, that protein, and all covariates, so the number of participants varies by protein depending on missingness. Across the 1,008 tests, the largest complete-case sample was 44,749 participants, and for AARSD1 (first protein in our results table) the complete-case sample was 44,016 participants (750 incident CAD cases and 43,266 controls). CAD was ascertained using UKB self-reported diagnosis codes (1075) and ICD-10 diagnosis codes (I210, I211, I212, I213, I214, I219, I220, I221, I228, I231,I232,I233,I236,I238, I241, I252). Each model regressed binary CAD status on a single standardized protein level, adjusted for age, sex, glycemic status (normoglycemia, prediabetes, incident T2D), assessment center, household income, and 40 genetic principal components. Proteins were tested individually to obtain interpretable per-protein effect estimates (odds ratios), each representing the association between a one-standard-deviation increase in protein level and incident CAD. Benjamini-Hochberg correction^65^ was applied across all 1,008 tests, with significance defined at FDR < 0.05.

#### Pharmacologic targetability of proteins

To assess pharmacologic targetability of proteins identified through MAP-D, we systematically queried the DrugBank database^27^ for approved drugs with documented protein targets. For each drug entry, we extracted identifiers (DrugBank ID, drug name), approval status, and associated target information, including UniProt accession, gene symbol, and protein name. Action types (e.g., inhibitor, antagonist, agonist, activator, stimulator, blocker, suppressor) were captured when annotated as “known” interactions in the database. Proteins significantly associated with cardiometabolic outcomes in MAP-D (FDR < 0.05) were matched to DrugBank targets using both UniProt IDs and gene symbols to maximize coverage. To ensure biological relevance, we restricted findings to approved drugs with directionally concordant actions. Specifically, proteins positively associated with disease risk (higher protein abundance associated with higher odds of incident CAD) were matched to inhibitors, antagonists, or blockers, while proteins inversely associated with disease risk were matched to agonists or activators. Drug–protein pairs were prioritized if they were directionally matched and associated with incident outcomes at FDR < 0.05. For summary statistics, we reported the number of unique drug–protein pairs, unique drugs, and unique proteins for CAD. Non-oncology agents were highlighted to emphasize clinical translatability and repurposing potential.

### QUANTIFICATION AND STATISTICAL ANALYSIS

All statistical analyses were performed in R (version 3.6.1). Multiple testing correction for proteome-wide associations used Bonferroni correction at p < 0.05/26,307. Mendelian randomization used IVW as the primary method with MR-Egger sensitivity analysis; Cochran’s Q assessed heterogeneity. Incident CAD associations applied Benjamini-Hochberg correction at FDR < 0.05 across 1,008 tests. GWAS were conducted using REGENIE v4.1.2.

## Supplemental Figures

**Supplemental Figure S1:**
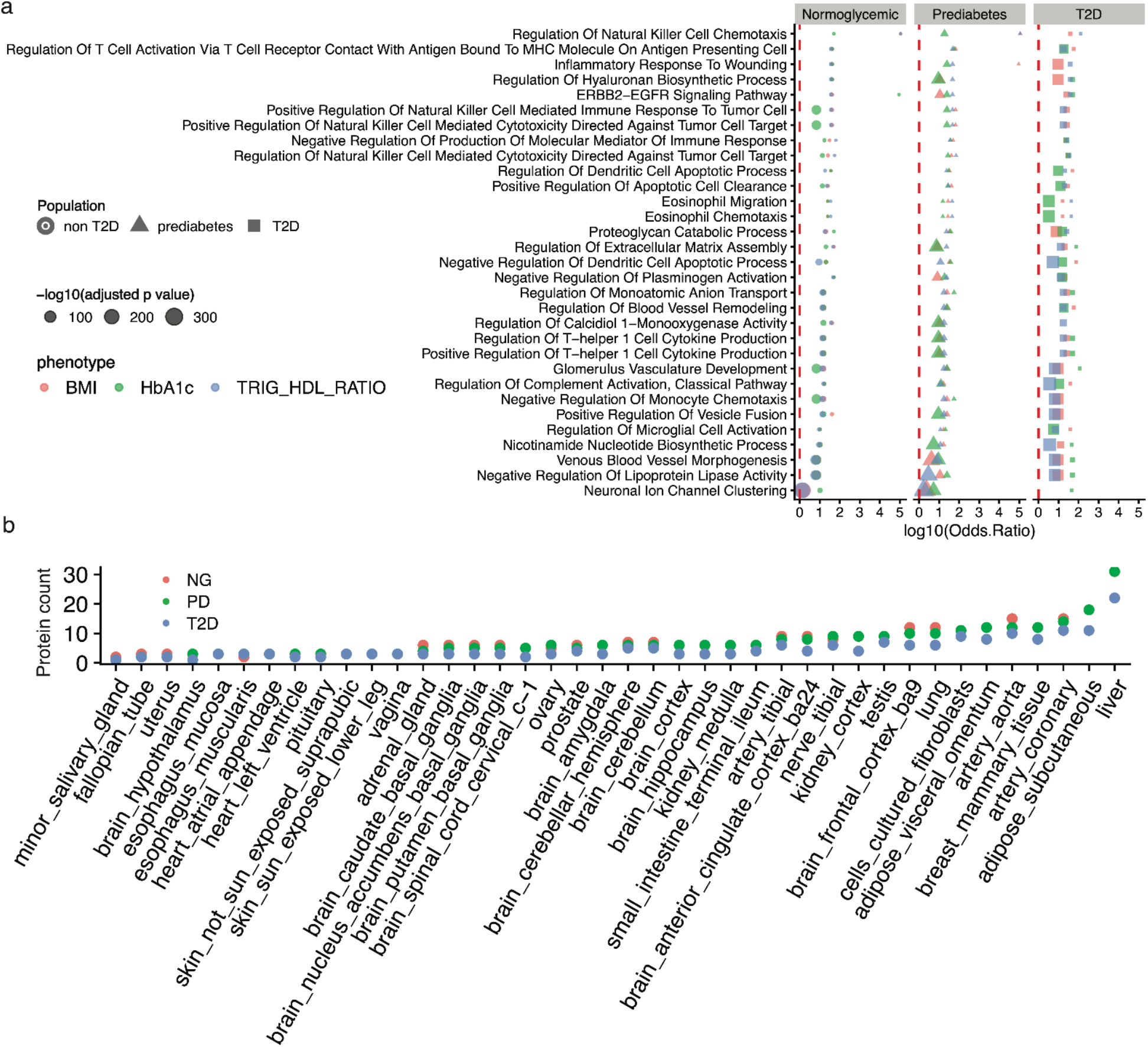
Comparing proteomic association across hallmarks. A) Dot plot of top enriched pathways across traits and glycemic stages, with point size indicating statistical significance (−log_10_[FDR corrected p-value]) and color denoting trait. B) Bar plot of pairwise correlations between hallmark indicators stratified by glycemic status. C) UpSet plot depicting number of shared protein associations across hallmark indicator and glycemic status combinations.

**Supplemental Figure S2:**
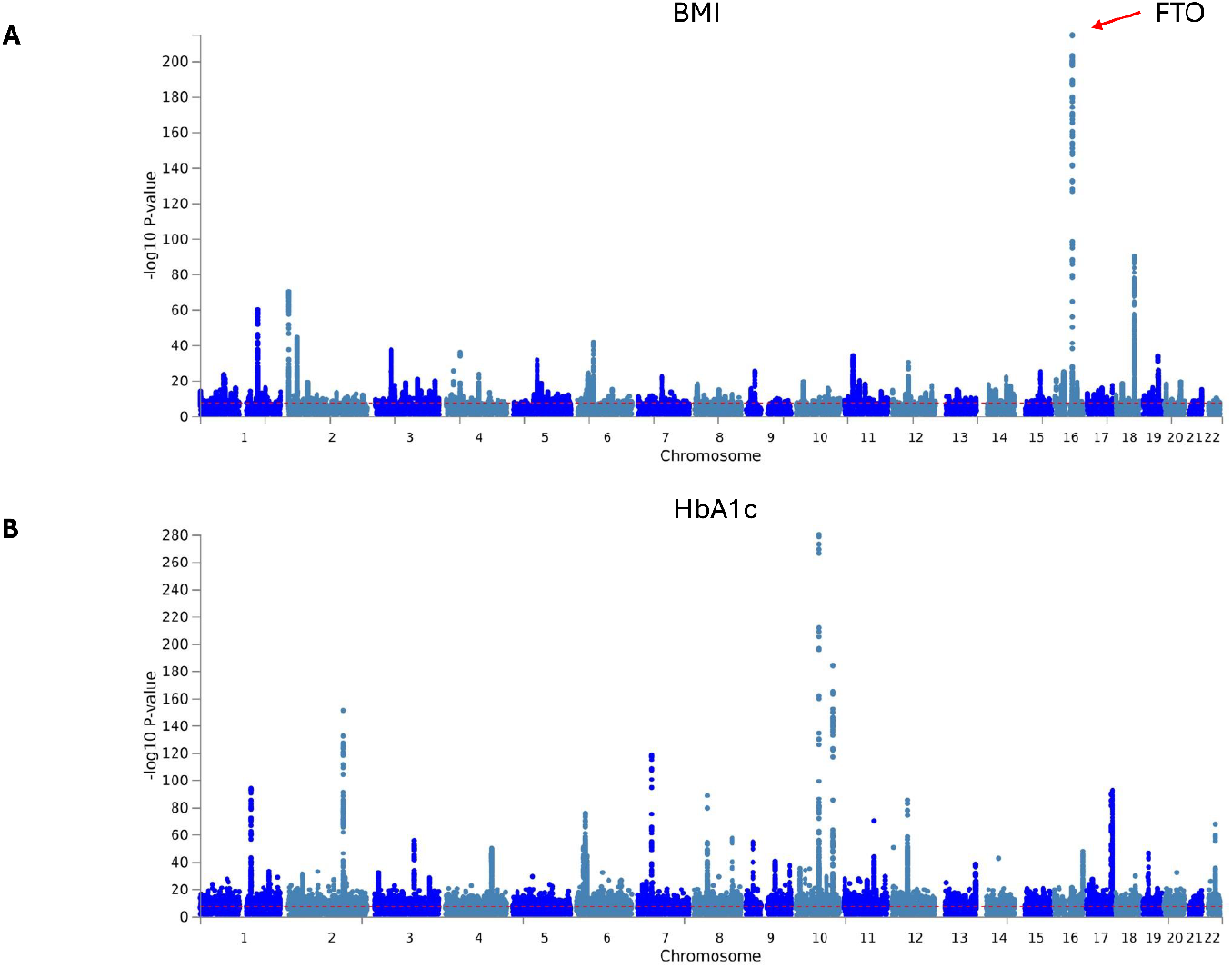
Genome-wide association of BMI and HbA1c in the UK Biobank held-out sample. A) Manhattan plot for body mass index (BMI) from REGENIE GWAS in participants in the full glycemic stratum (n ≈ 418,000 held-out non-proteomics individuals; Methods). The red horizontal dashed line indicates genome-wide significance (p-value = 5 x 10^-8^). B) Manhattan plot for glycated hemoglobin (HbA1c) under the same design. Chromosomes are shown on alternating colours along the x axis; the y axis shows −log_10_(p-value).

**Supplemental Figure S3:**
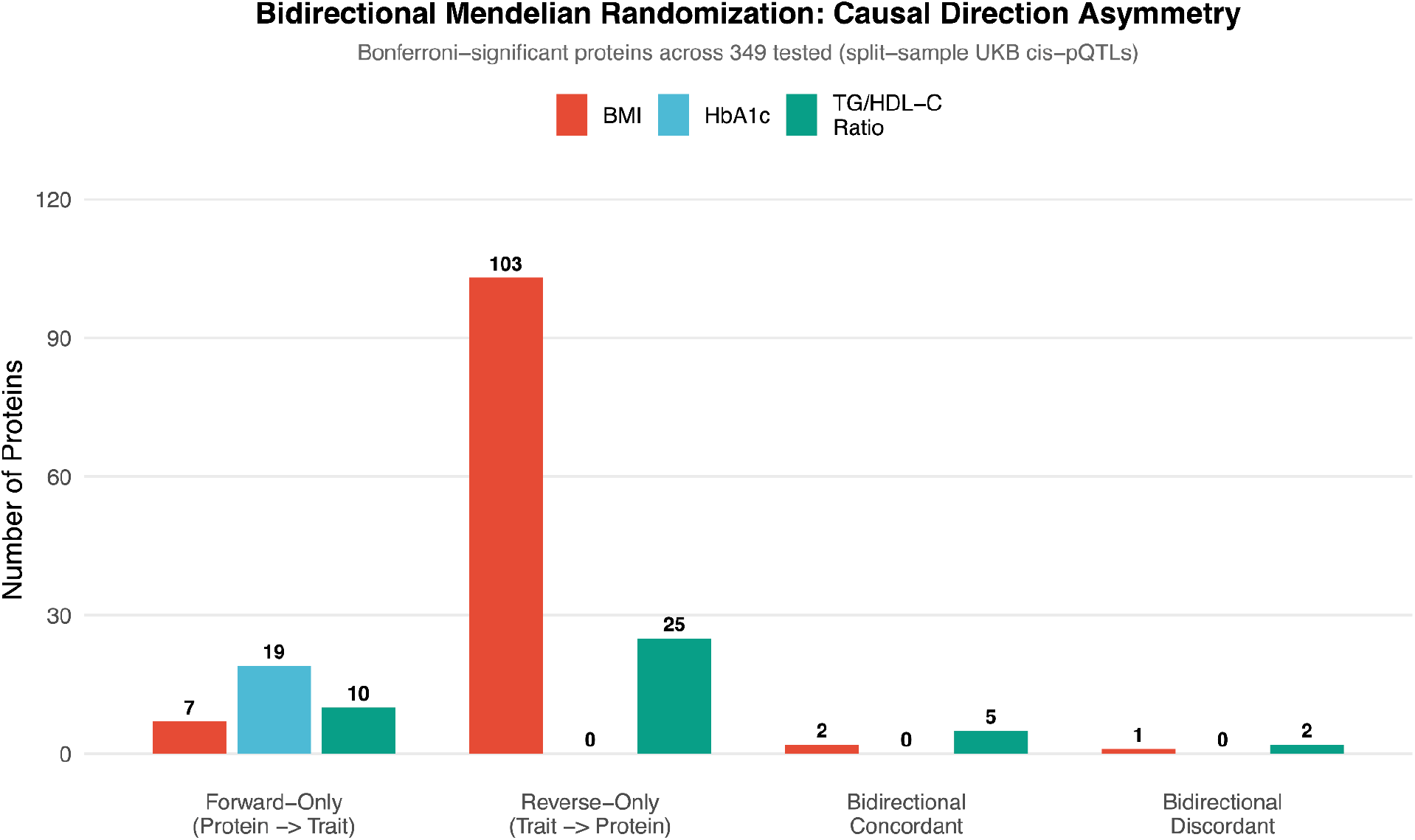
Causal direction asymmetry across BMI, HbA1c, and TG/HDL-C. Bar plot of protein associations stratified by direction for each hallmark.

**Supplemental Figure S4.**
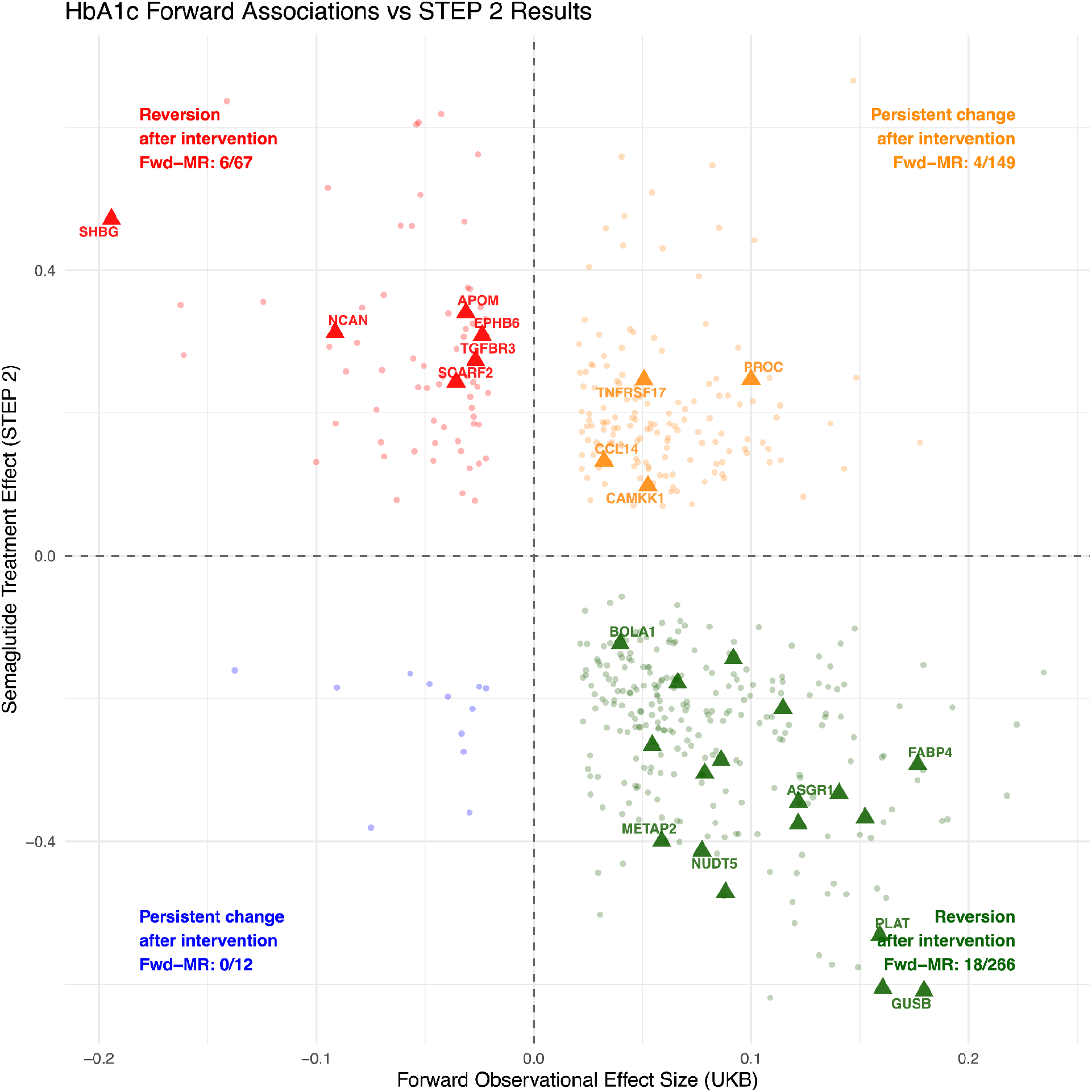
Comparing protein-first HbA1c associations with post-treatment change in STEP 2. Quadrants classify proteins by concordant or discordant directionality: reversion (red, green) or persistence (orange, blue). Red denotes UKB-/GLP-1RA+; green denotes UKB+/GLP-1RA-; yellow: UKB+/GLP-1RA+; blue: UKB-;GLP-1RA-. Square indicates a significant protein-first (protein → hallmark) MR association **concordant** with direction of the UKB and STEP trial. ETD: Estimated Treatment Difference.

**Supplemental Figure S5.**
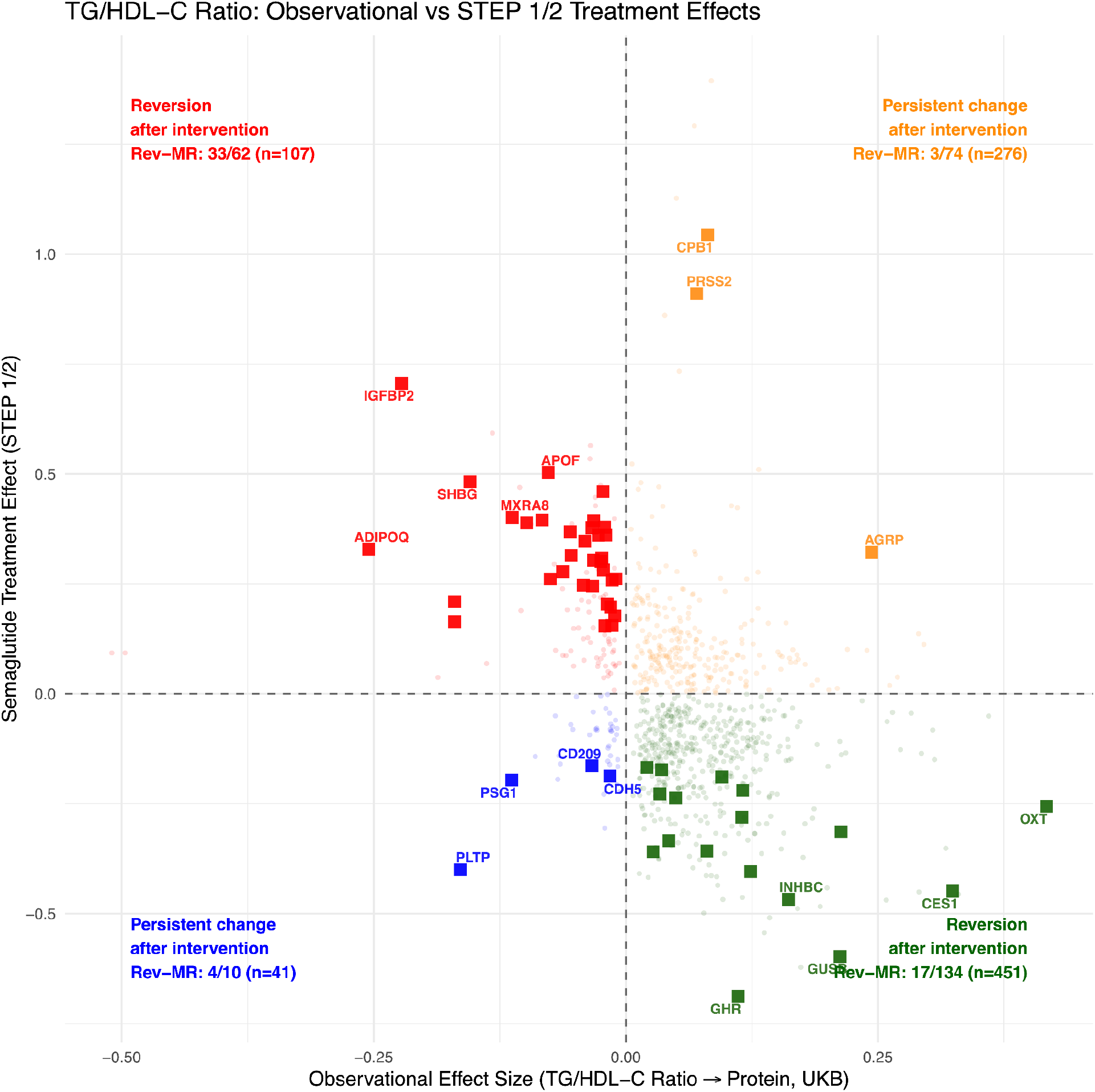
Comparing hallmark-firstTG/HDL-c associations with post-treatment change in STEP 1/2. Quadrants classify proteins by concordant or discordant directionality: reversion (red, green) or persistence (orange, blue). Red denotes UKB-/GLP-1RA+; green denotes UKB+/GLP-1RA-; yellow: UKB+/GLP-1RA+; blue: UKB-;GLP-1RA-. Square indicates a significant hallmark-first (hallmark → protein) MR association **concordant** with direction of the UKB and STEP trial. ETD: Estimated Treatment Difference.

## Notes

### Competing Interest Statement

The authors have declared no competing interest.

### Author Declarations

The data from the UK Biobank that support the findings of this study are available upon application (https://www.ukbiobank.ac.uk/register-apply/). Code for this manuscript is available via Zenodo at https://doi.org/10.5281/zenodo.1707108755 and https://github.com/stejat98/MAP-D. The MAP-D atlas data are available at https://doi.org/10.6084/m9.figshare.30007306 56. Additionally we make the GWAS summary statistics for the BMI, HbA1c, and TG/HDL Ratio hallmarks available at: https://doi.org/10.6084/m9.figshare.32048550 57. An interactive app can be found here: https://mapd.bio and https://chiragjp-mapd.share.connect.posit.cloud/.

### Summary of Updates

We added genetic associations to bolster causal claims. We updated the figures for clarity.

